# SOCIAL INEQUITIES IN A PSYCHOLOGICAL DOMAIN OF FOOD INSECURITY AMONG MOTHERS FROM SOUTHERN BRAZIL DURING THE COVID-19 PANDEMIC

**DOI:** 10.1101/2022.08.26.22279279

**Authors:** Thais Martins-Silva, Marina Xavier Carpena, Cauane Blumenberg, Rafaela Costa Martins, Kamyla M. Olazo, Bianca Del-Ponte, Luana P. Marmitt, Rodrigo Meucci, Juraci A. Cesar, Angela C. B. Trude, Christian Loret de Mola

## Abstract

We aimed to investigate the social inequalities in the fear of not having enough food for the household, a psychological domain of food insecurity, during the COVID-19 pandemic among mothers from the 2019 Rio Grande birth cohort. In 2019 we invited all mothers who gave birth to a singleton liveborn in the municipality of Rio Grande (southern Brazil) to respond to a standardized questionnaire. In 2020, we followed these mothers twice. In wave I we located 1,077 of all eligible mothers and 1,033 in wave II (follow-up rate of 52.1% and 50.4%). We estimated the absolute and relative inequalities of food insecurity according to the number of people living in the household, maternal education, family income, and income change during the pandemic using the slope index of inequalities (SII) and the concentration index (CIX), respectively. 1,021 mothers reported having food insecurity, and the prevalence was 42.8% and 44.7% for waves I and II, respectively. In wave I we observed inequities in the distribution of food insecurity, being higher among mothers living with ≥3 people (SII:-17.3; 95%CI[-29.6;-5.0]), with a lower education (SII:-36.7; 95%CI[-47.0;-26.4]), lower income (SII-48.9; 95%CI[-58.0;-39.7]), and with an income that decreased during the pandemic (SII:-47.1; 95%CI[-57.3;-36.9]). These inequities widened from wave I to wave II. This study describes the impact of the COVID-19 pandemic on maternal and family food insecurity and the increased social disparities during the pandemic, especially among the most vulnerable populations.

## INTRODUCTION

The new Coronavirus (COVID-19) was declared a pandemic by the World Health Organization on March 11, 2020, with social distancing measures starting in late March in most Brazilian cities. The social and economic responses to the pandemic (e.g., school and commerce closures, stay-at-home orders, and reduced government welfare coverage) resulted in unemployment, which was reflected in a decline in household purchasing power, and a broken food system due to losses in the supply of fresh foods, limited food transportation, and increased food prices (1). Therefore, the pandemic expanded food insecurity and its health-related disparities worldwide when comparing the most vulnerable to the better-off populations (2,3). Maternal and child mortality and morbidity increased during the pandemic, not only directly from COVID-19 but also due to indirect causes such as related illnesses and economic consequences that might critically influence the lack of resources (4). Limited financial resources and economic breakdown experienced by many families contributed to the increase in the rates of food insecurity, due to the lack of consistent physical, social, and economic access to adequate and nutritious food meeting dietary needs and food preferences (5). This collapse in food security can lead to serious public health consequences (6,7) including poor diet quality (8,9) and malnutrition (10).

The Global Report on Food Crises (11) estimates that 135 million people had food insecurity in 2019, and the World Food Program projects that this number could almost double, reaching 265 million people (12). However, little is known about how food and nutritional security were affected by the social and economic impacts of COVID-19, especially if pre-existing social inequities regarding income, ethnicity, gender, and access to health services are considered (1). In low- and middle-income countries with great social inequalities, such as Brazil (13), the social distancing measures adopted to curb the spread of the COVID-19 could further aggravate the food insecurity of certain population subgroups (1). For instance, it was estimated that approximately one-third of the urban households of Rio Grande (a Southern Brazilian city) experienced food insecurity in 2016 (14).

Worrying about food (psychological distress) is one of the earliest indicators of food insecurity (15). Therefore, the determinants of food insecurity must not only be framed by combined measures but also disaggregated according to subgroups of the population to support the implementation of effective public policies.

This study aimed to investigate the social inequalities in the fear of not having enough food for the household, a psychological domain of food insecurity, during the COVID-19 pandemic among mothers from the 2019 Rio Grande birth cohort.

## MATERIAL AND METHODS

### Participants

We used data from the WebCovid-19 study, a web-based follow-up of the 2019 Rio Grande birth cohort, an ongoing longitudinal study conducted in the municipality of Rio Grande (Southern Brazil) (16). Between January 1^st^ and December 31^st^ of 2019 (baseline), all hospital births in the city were identified (n=2,051) and mothers who gave birth to liveborn singletons and lived in the city’s urban area were invited to enroll in the birth cohort. In total, 2,314 out of 2,365 women, accepted to participate. A standardized questionnaire was applied by trained interviewers up to 48 hours after delivery. We collected general information including sociodemographic characteristics, habits, health status, place of residence, and cohabitants, among others. Between May and July 2020 (wave I of data collection), and July and December 2020 (wave II of data collection), all mothers who gave birth at baseline to a liveborn and lived in the city’s urban area (n=2,051) were invited to participate in the WebCovid-19 online follow-ups. They were contacted via phone calls, WhatsApp, or Facebook messages to answer a self-reported web-based questionnaire as part of the WebCovid-19 study. The REDCap software was developed and managed the web-based survey (17).

### Variables

Uncertainty and worry about the lack of food, a psychological domain of food insecurity (15), was considered the outcome of this study and is hereafter referred to as “food insecurity” for simplicity. Food insecurity was self-reported by mothers and collected during both waves of the WebCovid-19 study with the sentence: *“I am concerned about not having enough food, milk or other essentials for my baby”* (response options were yes or no). Mothers were asked to report if this statement applied to them in the last seven days before the survey.

To assess inequality dimensions, the following socio-economic and demographic variables were collected at baseline: the number of people in the household (<3; 3 or 4; or 5 or more); maternal schooling (elementary school; high school; or college or more), family income (in Brazilian *Reais – BRL –* and classified in tertiles), and self-reported changes in family income pre- and during the COVID-19 pandemic periods (increased, decreased, or stable).

### Statistical analysis

We calculated absolute and relative frequencies, and their 95% confidence intervals (95% CI). Chi-squared heterogeneity tests were used to calculate p-values between participants who were followed in both waves and those who were not. The prevalence of food insecurity was described according to the levels of the same variables.

Absolute and relative inequalities were calculated by estimating the slope index of inequalities (SII) and the concentration index (CIX), respectively. The SII was calculated by logistic regression and depicts the absolute difference (expressed in percentage points) in the prevalence of the outcome between the top and bottom of the distribution of the socioeconomic and demographic variables considered as inequality dimension. For instance, considering family income, the SII represents the absolute difference of the outcome, in percentage points, between the richest and poorest tertiles. In turn, the CIX is an indicator that depicts the relative difference in the prevalence of the outcome according to the levels of socioeconomic and demographic characteristics. CIX is related to the Gini coefficient, showing how much of the outcome is concentrated in the richest group compared to the poorest group. Zero values for both SII and CIX represent the absence of inequality; positive values indicate a higher prevalence of the outcome in the better-off group, and negative values indicate a higher prevalence in the worst-off group. These indexes have been extensively used in the inequity literature (18). Equiplot graphs were used to better illustrate the differences between the subgroups of the inequality dimensions. Differences in the prevalence of the outcome according to the socioeconomic and demographic variables were compared using Pearson’s chi-squared tests in each wave of data collection. All statistical analyses were performed using the Stata software version 16.1 (Stata Corporation, College Station, TX, USA) and considered a significance level of 5%.

### Ethics

The study was approved by the Ethics Committee of the *Universidade Federal do the Rio Grande*, under protocol number 15724819.6.0000.5324. An informed consent form was signed by all participants during the baseline study. For WebCovid-19 follow-up, a digital acceptance of terms and conditions was obtained from the mothers before the start of the survey.

## RESULTS

We located 1,077 (52.1%) and 1,033 (50.4%) of all eligible mothers in waves I and II, respectively. For this analysis, we included 1,021 mothers with complete information on food insecurity in wave I and 996 mothers in wave II (of a total of 1,297 individuals). The characteristics of these mothers are shown in Table 1. The prevalence of food insecurity was 42.8% and 44.7% for waves I and II, respectively. According to Supplementary Table S1, the participants included in our analyses were different from those not included due to losses to follow-up or missing information in the three inequality dimensions (number of people in the household, maternal schooling, and family income) (p<0.001).

**Table 1.**
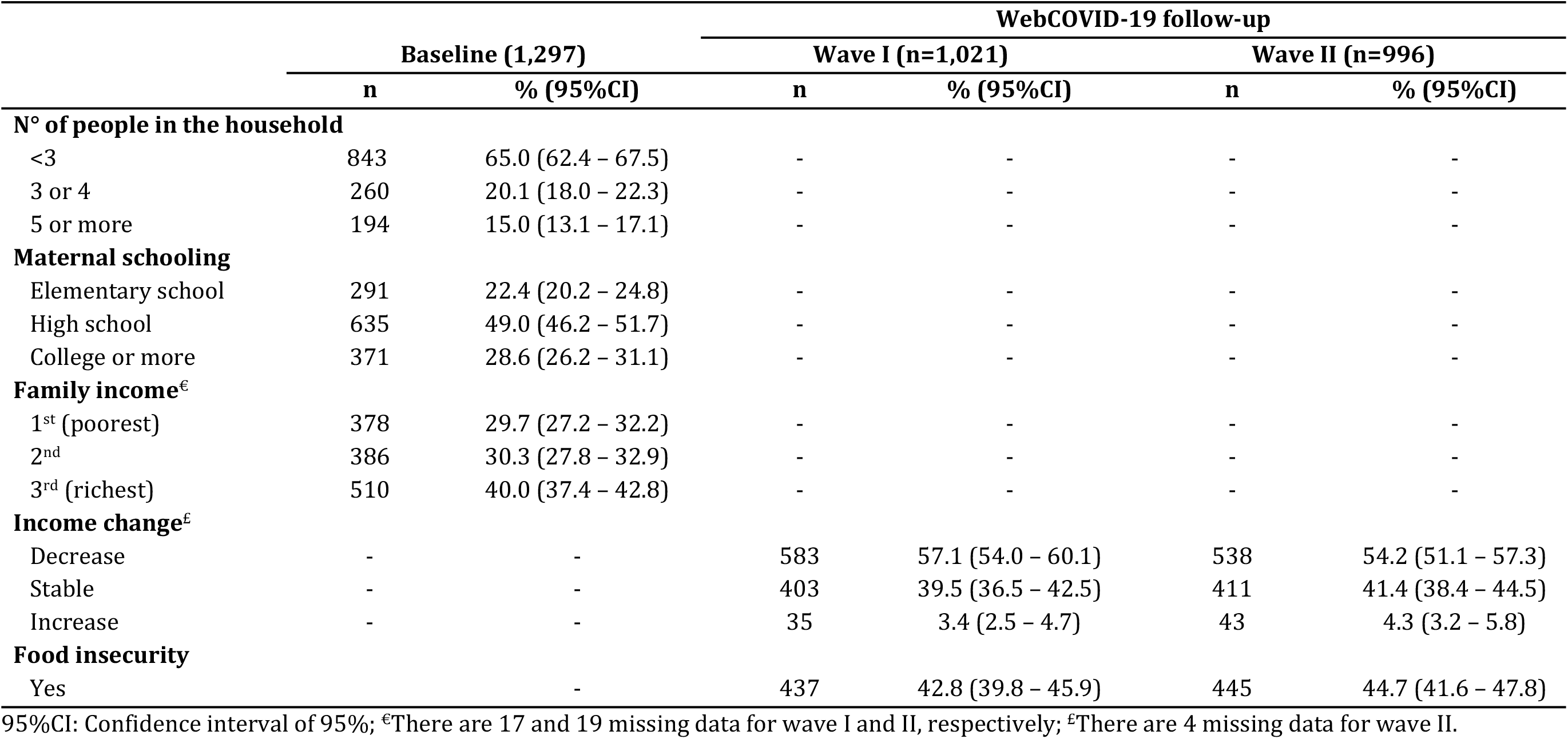
Sample description of baseline and WebCOVID-19 follow-up. 2019 Rio Grande Birth Cohort.

In wave I, the highest prevalence of food insecurity was found among mothers living with 3 or 4 people in the household (53.6%), who had an elementary school education (55.6%) and reported a reduction in family income between the pre- and during COVID-19 pandemic periods (53.9%) (Table 2). In wave II, the prevalence of food insecurity was higher among mothers living with 5 or more people in the household (52.7%), with an elementary school education (60.7%), and who reported a decrease in family income between the pre- and during the COVID-19 pandemic periods (59.1%). In addition, the prevalence of food insecurity according to family income was 2-fold higher among the poorest group compared to the richest in both waves (60.1% and 65.3% for waves I and II, respectively).

**Table 2.** Descriptive analysis of food insecurity and absolute (Slope Index of Inequality, SII) and relative (Concentration Index, CXI) inequalities among mothers including in the WebCOVID-19 follow-up, 2019 Rio Grande Birth Cohort.

| Variables related to inequalities dimensions | Food insecurity<sup>‡</sup> |  |  |  |  |  |
| --- | --- | --- | --- | --- | --- | --- |
|  | Wave I |  |  | Wave II |  |  |
|  | % (95%CI) | SII (95%CI) | CIX (95%CI) | % (95%CI) | SII (95%CI) | CIX (95%CI) |
| **N° of people in the household** |  | 17.0 (4.9;29.0) | 3.5 (-0.6;7.6) |  | 22.0 (10.0;33.9) | 5.5 (1.5;9.4) |
| <3 | 39.5 (35.9; 43.2) |  |  | 40.6 (36.9; 44.4) |  |  |
| 3 or 4 | 53.6 (46.6; 60.4) |  |  | 52.4 (45.3; 59.4) |  |  |
| 5 or more | 44.1 (36.0; 52.6) |  |  | 52.7 (44.7; 60.5) |  |  |
| **Maternal schooling** |  | -36.0 (-46.1;-25.9) | -12.7 (-16.7;-8.7) |  | -42.7 (-52.5;-32.9) | -14.0 (-17.9;-10.1) |
| Elementary school | 55.6 (48.7; 62.3) |  |  | 60.7 (53.9; 67.0) |  |  |
| High school | 46.6 (42.2; 51.0) |  |  | 48.2 (43.8; 52.7) |  |  |
| College or more | 28.8 (24.1; 34.0) |  |  | 28.1 (23.3; 33.4) |  |  |
| **Family income<sup>€</sup>** |  | -48.0 (-57.0;-39.0) | -16.9 (-20.9;-12.8) |  | -54.1 (-62.6;-45.5) | -18.7 (-22.7;-14.8) |
| 1<sup>st</sup> (poorest) | 60.1 (54.2; 65.8) |  |  | 65.3 (59.5; 70.6) |  |  |
| 2<sup>nd</sup> | 48.5 (42.9; 54.2) |  |  | 49.3 (43.5; 55.1) |  |  |
| 3<sup>rd</sup> (richest) | 27.2 (23.2; 31.6) |  |  | 27.3 (23.2; 31.9) |  |  |
| **Income change<sup>£</sup>** |  | -46.2 (-56.3;-36.1) | -15.0 (-19.1; -11.0) |  | -55.9 (-64.9;-46.9) | -17.6 (-21.6;-13.7) |
| Decrease | 53.9 (49.8; 57.9) |  |  | 59.1 (54.9; 63.2) |  |  |
| Stable | 27.1 (22.9; 31.6) |  |  | 27.0 (22.9; 31.5) |  |  |
| Increase | 40.0 (25.3; 56.7) |  |  | 30.2 (18.4; 45.4) |  |  |

95%CI: Confidence interval of 95%; SII: Slope Index of Inequality expressed in percentage points; CIX: Concentration Index (x 100)
<sup>‡</sup>Data presented for the category "yes" of each variable; <sup>€</sup>There are 17 and 19 missing data for wave I and II, respectively; <sup>£</sup>There are 4 missing data for wave II.

Table 2 shows that food insecurity was also higher among mothers who lived with <3 people in the same household (SII:17.0; 95% confidence interval [CI][4.9;29.0]), had lower maternal schooling (SII:-36.0; 95%CI[-46.1;-25.9]), belonged to the poorest tertile of family income (SII:-48.0; 95%CI[-57.0;-39.0]), and had a reduction in family income during wave I of the COVID-19 pandemic (SII:-46.2; 95%CI[-56.3;-36.1]). In wave II these indicators were even higher, suggesting that inequities increased from wave I to wave II. However, there was an overlapping between the confidence intervals.

Figure 1 is an equiplot showing the prevalence of food insecurity in each category of schooling and income at baseline, and changes in income during the pandemic. The distance between the dots represents prevalence differences and inequity in the distribution. This figure shows that not only the poorer, the less educated, and those with a decreased income had the highest prevalence, but the distribution was unequal among the categories. The distance for those in the highest categories was higher, suggesting an uneven distribution of food insecurity. The burden is concentrated among the less educated and with less income.

**Figure 1.**
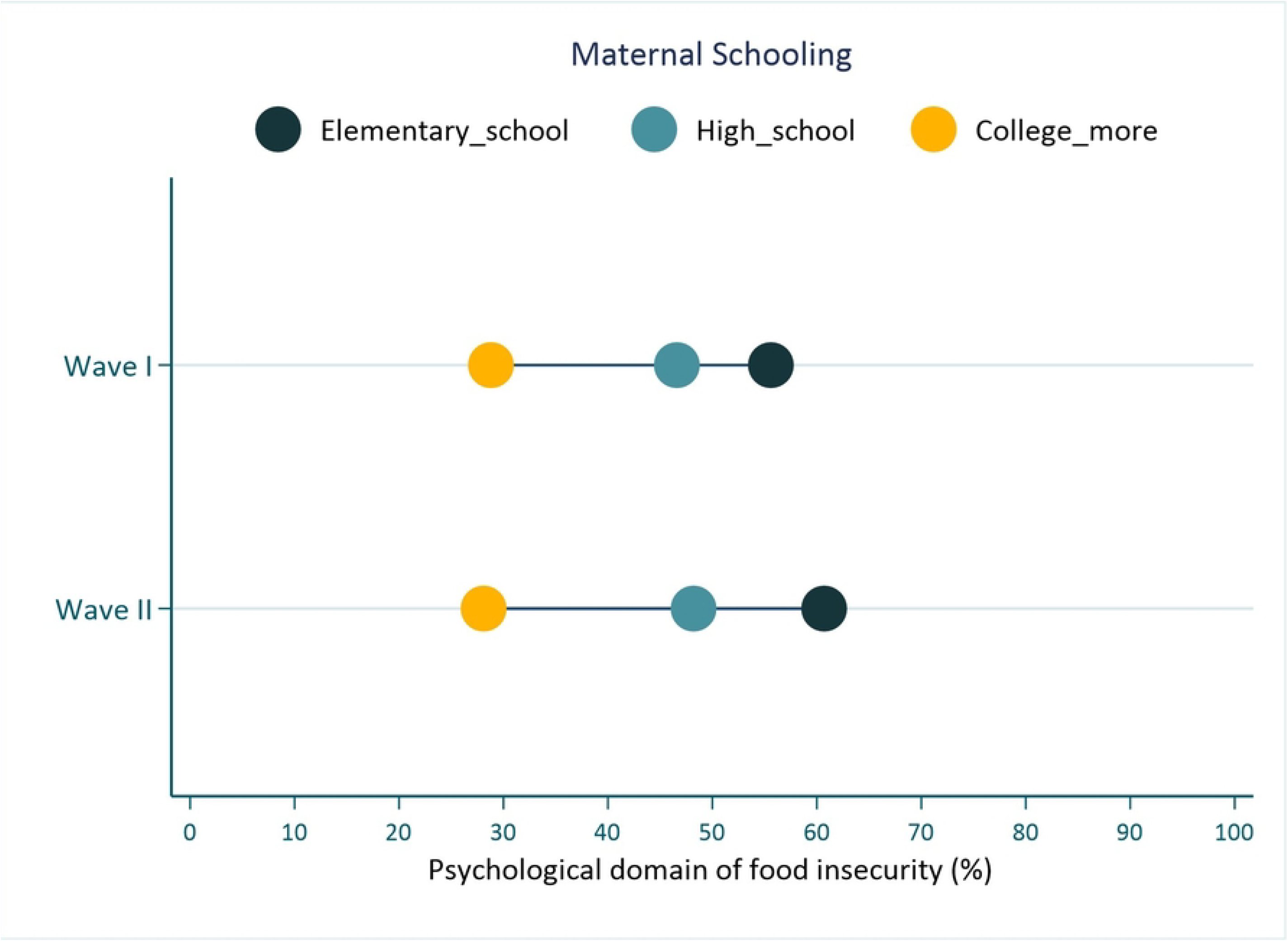
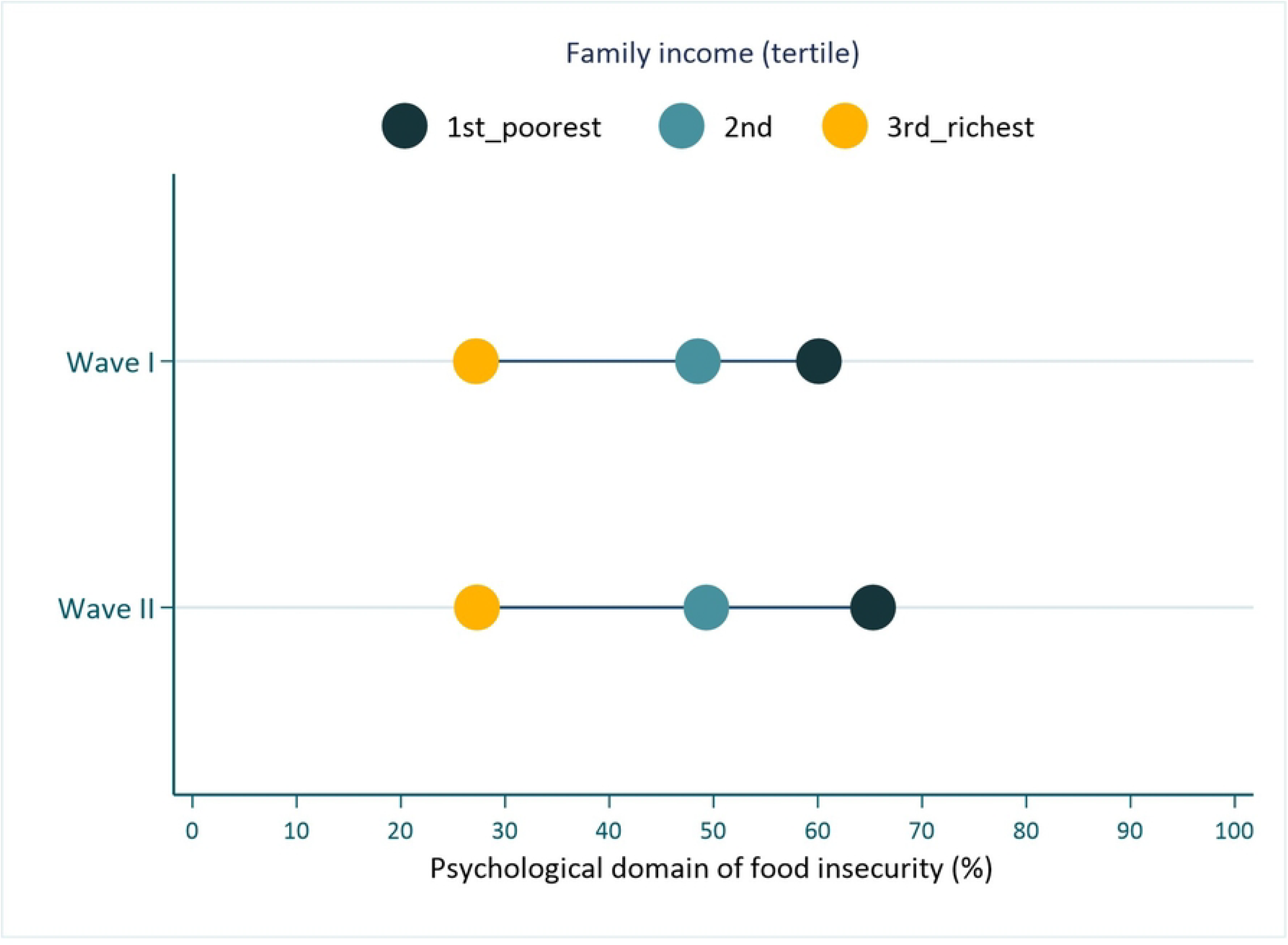
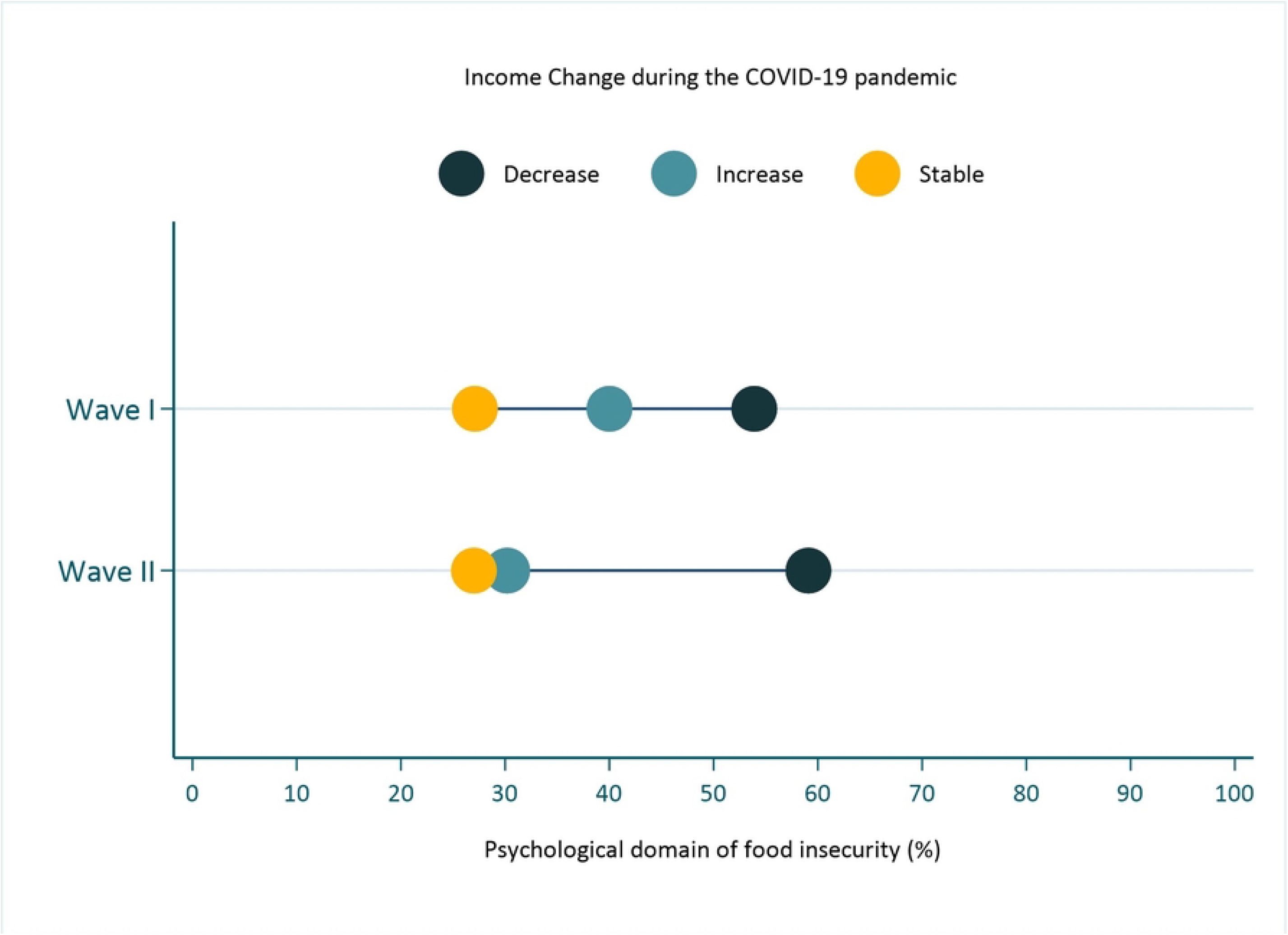
Prevalence of psychological domain of food insecurity according to (a) maternal schooling, (b) family income, and (c) income change during COVID-19 pandemic among mothers including in the WebCOVID-19 follow-up, 2019 Rio Grande Birth Cohort.

## DISCUSSION

The findings of this study show that the prevalence of food insecurity (uncertainty and worry about food) during the first year of the pandemic was higher than what was previously reported for this population (14). It is alarming to see that more than half of the mothers of the city with a child of less than two years of age reported being concerned about not having enough food, milk, or other essentials for their babies and that this insecurity was concentrated in families who had fewer economic resources, a reduction in income during the pandemic, among mothers with a lower education, and in households with a higher number of individuals.

Regarding the prevalence of food insecurity, our results are consistent with previous studies that demonstrate that food insecurity increases during socioeconomic collapse among the most vulnerable households (19,20) and that, as the pandemic advances, food security can worsen even further (21,22). This rise in food insecurity among mothers has many negative health consequences, which are associated with increased risks of birth defects, anemia, cognitive problems, aggression, and anxiety (23). It is also associated with higher risks of being hospitalized and poorer general health and with having asthma (24), worse oral health (25–28), and poor diet quality (29,30). In the present study, mothers experiencing food insecurity demonstrated significantly higher rates of concern and worry about food. Food and nutritional insecurity and/or lower socio-economic levels are associated with greater consumption of food poor in nutrients and with high energy density, which generally have a lower cost (31) and are associated with morbidities, such as obesity, hypertension, and type 2 diabetes mellitus (32).

The COVID-19 pandemic has sparked fear and concern in many countries not only because of the virus itself but also due to the restrictive public health measures implemented to reduce community transmission (33). This is likely to increase stress and anxiety indicators (16), particularly among mothers who already have a level of concern for their infants. However, the pandemic also caused an intensification of women’s daily routines, including childcare, house cleaning, and meals preparation, in addition to the increase in domestic violence and the contingent social distancing during the pandemic (34), which had a direct impact on the general health of this population. Moreover, family income may contribute to the quality of nutrition (35). Further research is needed to understand how food insecurity during the COVID-19 pandemic is related to diet quality, particularly if disrupted eating patterns persist and increase.

In Brazil, the COVID-19 pandemic exposed multiple vulnerabilities that had already been festering during the years previous to the pandemic. Although a trending decrease in food insecurity was observed in Brazil from 2004 to 2013 (17.0% to 7.9%, respectively), this condition has always been marked by great inequalities (36). A higher prevalence of food insecurity was observed among the poorest, those living in urban areas with inadequate sanitation, overcrowded housing, female-headed households, individuals younger than 60 years of age, non-whites, having less than four years of schooling, and being unemployed (21). Even though this prevalence has declined, it was largely concentrated among these subgroups of the population over the years (2004-2013), and these same groups are also more likely to be affected by the dissemination of COVID-19 and its consequences (36).

It is true that the social risk factors for food insecurity in Brazil were similar before and during the COVID-19 pandemic; however, the increase in the prevalence of food insecurity has dramatically magnified the burden and consequences on the most vulnerable and should be of great concern (37).

Our results show that the prevalence of food insecurity increased especially among the poorest families, potentially widening social inequalities during the COVID-19 pandemic. Moreover, the closure of commercial activities and other economic sectors due to the pandemic in Brazil had a negative consequence on the economy (7). A comparison between the final quarter of 2019 and the first quarter of 2020 revealed an increase in the unemployment rate, the highest in the Northeastern region, raising the number of poor people in Brazil (38). By June 2020, the job contracts or working hours and salaries of more than 10 million workers had already been suspended or reduced. The increase in unemployment and poverty in the country combined with inflation, (39) further increased the number of Brazilians that are vulnerable to food and nutrition insecurity.

Therefore, although there was no interruption in food supply, Brazilians with compromised incomes during the pandemic may have had problems in accessing food because they were unable to afford it. Given the negative impacts caused by the COVID-19 pandemic on food insecurity (21), the Brazilian government responded with some, albeit slow and uncoordinated, actions to minimize the harm of the pandemic to the most vulnerable families. For instance, an emergency aid payment of US$120 was distributed over 5 months to informal workers, individual entrepreneurs, independent workers, and the unemployed. By August 2020, 66.2 million Brazilians had already received this payment. However, the management and distribution of this aid has been heavily criticized due to the difficulties and delays during the process which further exacerbated the feelings of insecurity during a political crisis (39).

Another issue that increased food insecurity during the COVID-19 pandemic was the closure of schools, which impacted the distribution of food, especially to the poorest children. Brazil is one of the top countries in implementing school feeding programs, and families that had already struggled to make ends meet also had to manage an increase in household food expenditure due to the number of meals per individual in the household. Thus, school closure can increase the probability of food insecurity and have a negative impact on children’s health and development (40).

This study has some limitations. First, almost half of the participants did not participate in the follow-ups, and therefore, the findings of this study should be interpreted in light of this limitation. In the current study, most of the people not included were of the poorest family income, had less maternal schooling, and lived with more people in the household. Taking this into account, food insecurity may have been underestimated, and its inequalities are most likely greater than what is shown in our analysis.

Our findings highlight the need for policies and interventions to reduce the impact of the COVID-19 pandemic on maternal and family food insecurity and the increased social disparities during the pandemic, especially among the most vulnerable populations. A few potential strategies to mitigate these problems include extending the government COVID-19 assistance program, direct provision of food to families in need, and expanded access to healthy, affordable, and safe food.

## Data Availability

Data are available by the research group entitled "Grupo de Pesquisa e Inovação em Saúde (GPIS)" (contact via) for researchers who meet the criteria for access to confidential data.

## ACKNOWLEDGMENTS

This study was funded by Conselho Nacional de Desenvolvimento Científico e Tecnológico (CNPq), grant number 433426/2018-7, and the Rio Grande Municipal Department of Health. The study was conducted within a graduate program supported by the Coordenaçõa de Aperfeiçoamento de Pessoal de Nível Superior (CAPES; finance code 001). Moreover, this research was funded in whole, or part, by the Wellcome Trust [Grant number 210735_Z_18_Z]. For the purpose of open access, the author has applied for a CC BY public copyright license to any Author Accepted Manuscript version arising from this submission.

## SUPPORTING INFORMATION

**Supplementary Table S1**. Comparison of inequalities dimensions between those mothers included and not included in the current study for WebCOVID-19 follow-up waves I and II. 2019 Rio Grande Birth Cohort.

## REFERENCES

1. Ribeiro-Silva R de C, Pereira M, Campello T, Aragão É, Guimarães JM de M, Ferreira AJ, et al. Covid-19 pandemic implications for food and nutrition security in Brazil. Cien Saude Colet. 2020 Sep;25(9):3421–30.

2. Ahorsu DK, Lin CY, Imani V, Saffari M, Griffiths MD, Pakpour AH. The Fear of COVID-19 Scale: Development and Initial Validation. International journal of mental health and addiction. 2020. p. 1–9.

3. Pakpour AH, Griffiths M. The fear of COVID-19 and its role in preventive behaviours. Journal of Concurrent Disorders. 2020;

4. Kotlar B, Gerson E, Petrillo S, Langer A, Tiemeier H. The impact of the COVID-19 pandemic on maternal and perinatal health: a scoping review. Reprod Health. 2021;18(1):10.

5. Steimle S, Gassman-Pines A, Johnson AD, Hines CT, Ryan RM. Understanding patterns of food insecurity and family well-being amid the COVID-19 pandemic using daily surveys. Child Dev. 2021 Sep;92(5):e781–97.

6. Roberton T, Carter ED, Chou VB, Stegmuller AR, Jackson BD, Tam Y, et al. Early estimates of the indirect effects of the COVID-19 pandemic on maternal and child mortality in low-income and middle-income countries: a modelling study. Lancet Glob Health. 2020 Jul;8(7):e901–8.

7. Berkowitz SA, Basu S, Meigs JB, Seligman HK. Food Insecurity and Health Care Expenditures in the United States, 2011-2013. Health Serv Res. 2018 Jun;53(3):1600–20.

8. Wolfson JA, Leung CW. Food Insecurity and COVID-19: Disparities in Early Effects for US Adults. Nutrients. 2020 Jun 2;12(6):1648.

9. Leung CW, Epel ES, Ritchie LD, Crawford PB, Laraia BA. Food insecurity is inversely associated with diet quality of lower-income adults. J Acad Nutr Diet. 2014 Dec;114(12):1943-53.e2.

10. Ribeiro-Silva R de C, Pereira M, Aragão É, Guimarães JM de M, Ferreira AJF, Rocha A dos S, et al. COVID-19, Food Insecurity and Malnutrition: A Multiple Burden for Brazil. Front Nutr. 2021;8.

11. Global Network Against Food Crises; Food Security Information Network. Global Report on Food Crises. Joint analisys for better decisions. Washington DC: International Food Policy Research Institute; 2020.

12. Alpino T de MA, Santos CRB, Barros DC de, Freitas CM de. COVID-19 e (in)segurança alimentar e nutricional: ações do Governo Federal brasileiro na pandemia frente aos desmontes orçamentários e institucionais. Vol. 36, Cadernos de Saúde Pública. scielo; C de SP. scielo; 2020. No Title.

13. Pitombeira DF, de Oliveira LC. Pobreza e desigualdades sociais: tensões entre direitos, austeridade e suas implicações na atenção primária. Cien Saude Colet. 2020;25:1699–708.

14. Carvalho CA de, Viola PC de AF, Sperandio N. How is Brazil facing the crisis of Food and Nutrition Security during the COVID-19 pandemic? Public Health Nutr. 2021 Feb 12;24(3):561–4.

15. Johnson CM, Ammerman AS, Adair LS, Aiello AE, Flax VL, Elliott S, et al. The Four Domain Food Insecurity Scale (4D-FIS): development and evaluation of a complementary food insecurity measure. Transl Behav Med. 2020 Dec 1;10(6):1255–65.

16. Loret de Mola C, Martins-Silva T, Carpena MX, Del-Ponte B, Blumenberg C, Martins RC, et al. Maternal mental health before and during the COVID-19 pandemic in the 2019 Rio Grande birth cohort. Brazilian Journal of Psychiatry. scielo; 2021.

17. Harris PA, Taylor R, Thielke R, Payne J, Gonzalez N, Conde JG. Research electronic data capture (REDCap)—A metadata-driven methodology and workflow process for providing translational research informatics support. J Biomed Inform. 2009;42(2):377–81.

18. Barros AJD, Victora CG. Measuring coverage in MNCH: determining and interpreting inequalities in coverage of maternal, newborn, and child health interventions. PLoS Med. 2013;10(5):e1001390.

19. King C. Food insecurity and child behavior problems in fragile families. Econ Hum Biol. 2018 Feb;28:14–22.

20. Gundersen C, Kreider B, Pepper J. The Economics of Food Insecurity in the United States. Appl Econ Perspect Policy. 2011;33(3):281–303.

21. Manfrinato C V, Marino A, Condé VF, Franco M do CP, Stedefeldt E, Tomita LY. High prevalence of food insecurity, the adverse impact of COVID-19 in Brazilian favela. Public Health Nutr. 2021 Apr;24(6):1210–5.

22. Abay KA, Amare M, Tiberti L, Andam KS. COVID-19-Induced Disruptions of School Feeding Services Exacerbate Food Insecurity in Nigeria. J Nutr. 2021 Aug;151(8):2245–54.

23. Shankar P, Chung R, Frank DA. Association of Food Insecurity with Children’s Behavioral, Emotional, and Academic Outcomes: A Systematic Review. J Dev Behav Pediatr. 2017;38(2):135–50.

24. Clemens KK, L. B, Ouédraogo AM, Mackenzie C, Vinegar M, Shariff SZ. Childhood food insecurity and incident asthma: A population-based cohort study of children in Ontario, Canada. PLoS One. 2021;16(6):e0252301.

25. Chi DL, Masterson EE, Carle AC, Mancl LA, Coldwell SE. Socioeconomic Status, Food Security, and Dental Caries in US Children: Mediation Analyses of Data From the National Health and Nutrition Examination Survey, 2007–2008. Am J Public Health. 2014 May;104(5):860–4.

26. Howard LL. Does food insecurity at home affect non-cognitive performance at school? A longitudinal analysis of elementary student classroom behavior. Econ Educ Rev. 2011 Feb;30(1):157–76.

27. Carmichael SL, Yang W, Herring A, Abrams B, Shaw GM. Maternal Food Insecurity Is Associated with Increased Risk of Certain Birth Defects. J Nutr. 2007 Sep 1;137(9):2087–92.

28. Cook JT, Frank DA, Levenson SM, Neault NB, Heeren TC, Black MM, et al. Child Food Insecurity Increases Risks Posed by Household Food Insecurity to Young Children’s Health. J Nutr. 2006 Apr 1;136(4):1073–6.

29. André HP, Sperandio N, Siqueira RL de, Franceschini S do CC, Priore SE. Food and nutrition insecurity indicators associated with iron deficiency anemia in Brazilian children: a systematic review. Cien Saude Colet. 2018 Apr;23(4):1159–67.

30. Huet C, Rosol R, Egeland GM. The Prevalence of Food Insecurity Is High and the Diet Quality Poor in Inuit Communities. J Nutr. 2012 Mar 1;142(3):541–7.

31. Drewnowski A. The cost of US foods as related to their nutritive value. Am J Clin Nutr. 2010 Nov;92(5):1181–8.

32. Hamedi-Shahraki S, Mir F, Amirkhizi F. Food Insecurity and Cardiovascular Risk Factors among Iranian Women. Ecol Food Nutr. 2021;60(2):163–81.

33. Cowling BJ, Aiello AE. Public Health Measures to Slow Community Spread of Coronavirus Disease 2019. J Infect Dis. 2020 May 11;221(11):1749–51.

34. Marques ES, Moraes CL de, Hasselmann MH, Deslandes SF, Reichenheim ME. Violence against women, children, and adolescents during the COVID-19 pandemic: overview, contributing factors, and mitigating measures. Cad Saude Publica. 2020;36(4):e00074420.

35. Mello CS, Barros KV, de Morais MB. Brazilian infant and preschool children feeding: literature review. Jornal de Pediatria (Versão em Português). 2016;92(5):451–63.

36. Santos TG Dos, Silveira JAC da, Longo-Silva G, Ramires EKNM, Menezes RCE de. [Trends and factors associated with food insecurity in Brazil: the National Household Sample Survey, 2004, 2009, and 2013]. Cad Saude Publica. 2018;34(4):e00066917.

37. Santos, MPA, Nery, JS, Goes, EF et al. (2020) Black population and Covid-19: reflections on racism and health. Estud Av 34, 225–244.

38. Brazilian Institute of Geography and Statistics (2020) Continuous National Household Sample Survey-Continuous PNAD. https://www.ibge.gov.br/en/statistics/multi-domain/living-conditions-poverty-and-inequality/16809-quarterlydissemination-pnad2.html?edicao=27711&t=destaques (Accessed June 2022).

39. Silva Filho OJ da, Gomes Júnior NN. The future at the kitchen table: COVID-19 and the food supply. Cad Saude Publica. 2020;36(5):e00095220.

40. Mutisya M, Ngware MW, Kabiru CW, Kandala N bakwin. The effect of education on household food security in two informal urban settlements in Kenya: a longitudinal analysis. Food Secur. 2016;8(4):743–56.

